# Immune-oncology-microbiome axis may result in AKP or anti-AKP effects in intratumor microbiomes

**DOI:** 10.1101/2024.03.23.24304783

**Authors:** Zhanshan (Sam) Ma

**Affiliations:** Computational Biology and Medical Ecology Lab, Center for Excellence in Animal Evolution and Genetics, Chinese Academy of Sciences, Kunming, 650222, China; Microbiome Medicine and Advanced AI Lab, Cambridge, MA 02138, USA; Faculty of Arts and Sciences, Harvard University, Cambridge, MA 02138, USA

**Keywords:** AKP (Anna Karenina principle), Cancer tissue microbiome, TCGA (the cancer genome atlas), Cancer hallmark, Genome instability and mutation, Tumor microenvironment

## Abstract

An emerging consensus regarding the triangle relationship between tumor, immune cells, and microbiomes is the immune-oncology-microbiome (IOM) axis, which stipulates that microbiomes can act as a discrete enabling (or disabling) characteristic that broadly influence the acquisition of certain hallmarks of cancer, *i.e*., a set of functional capabilities acquired by human cells during carcinogenesis and progression to malignant tumors. Specifically, it has been postulated that polymorphic microbiomes can either induce or inhibit some of the hallmark capacities (particularly, immune evasions) via their intersecting with two other enabling characteristics (genome instability and mutation, and tumor promoting inflammation). The net effects of the microbiomes can be either protective or deleterious effects on cancer development, malignant progression, and therapy responses. Nevertheless, there is not yet a mechanistic interpretation for IOM, especially regarding intratumoral microbiomes. Here, we propose to interpret the observed relationships, in which microbiomes can be complicit, bystanders, or in rare cases, oncomicrobes or foes, to either cancer cells or immune cells, possibly depend on specific microbial taxon, with the AKP (Anna Karenina principle)—that all heathy tissue microbiomes should be similar, and tumor microbiomes should be dissimilar with each other, in analogy with Leo Tolstoy’s aphorism that “all happy families look alike; each unhappy family is unhappy in its own way”. Given potentially double-sword nature of microbes, both AKP and anti-AKP should exist in the IOM axis. We test the AKP with microbiome datasets of 20+ cancer types from the TCGA database and find that the ratio of AKP/anti-AKP is about 3:1.

**Key Points:**

- We propose to interpret the observed relationships, in which microbiomes can be complicit, bystanders, or in rare cases, oncomicrobes or foes, to either cancer cells or immune cells, possibly depend on specific microbial taxon, with the AKP (Anna Karenina principle).
- We postulate that all heathy tissue microbiomes should be similar, and tumor microbiomes should be dissimilar with each other, in analogy with Leo Tolstoy’s aphorism that “all happy families look alike; each unhappy family is unhappy in its own way”.
- We test the AKP with microbiome datasets of 20+ cancer types from the TCGA database and find that the ratio of AKP/anti-AKP is about 3:1.

## Introduction

At frontiers in microbiome research, it has been confirmed that all animals studied including humans, except for a couple of exceptions (Tobin *et al*. 2019, Ma *et al*. 2022), possess their own microbiomes, which play important, if not indispensable, roles in normal physiology (health) and susceptibility to disease. For this, predicting their responses to perturbations from internal or external is rather rewarding but challenging (Zaneveld *et al*. 2017). Treating diseases such as HIV and cancers as perturbations have been a well-received practice in the field of microbiome associated diseases.

Sepich-Poore *et al* (2021) introduced the concept of immune-oncology-microbiome (IOM) axis in their comprehensive review on the state-of-the-art research in the relationships between human cancers and microbiomes. Open questions in the field include: are microbes causal agents, complicit actors, or passive bystanders? And more importantly, what about intratumoral microbes? which we are particularly interested in this study. In the present study, we aim to investigate one aspect of the relationships, *i.e*., how would the intratumoral microbiome respond to the stress from cancer cells?

Among the estimated ∼10^12^ microbial species on the earth planet, only 11 species were labelled oncomicrobes or human carcinogens according to the IACR (international association for cancer registries) (Sepich-Poore *et al* (2021). Recent experimental evidence supports the notion of “complicit” microbes, who may promote carcinogenesis but insufficient to cause cancers (Sepich-Poore *et al*. 2021). To some extent, the complicit microbes are similar to opportunistic pathogens, rather than direct causal pathogens. The “complicit” category of microbes can produce bioactive metabolites, be responsible for many immunomodulatory functions of microbiome in tumor development, and linked to the immune system’s role in solid tumorigenesis (Sepich-Poore *et al*. 2021). Whether or not the complicit microbes are involved in cancer development may depend on their intersections with host genome instability and mutation and/or immune system modulations. For example, common p53 mutations are only carcinogenic in combination with microbially produced gallic acid and are even protective otherwise in the gut (cited in Sepich-Poore *et al*. 2021).

Concerning the relationships between human microbiomes and cancers, there are at least three levels (scales) to pursue their investigations: (*i*) the gut (intestinal) microbiome and cancers including extraintestinal cancers; (*ii*) local microbiomes in extraintestinal cancers such as respiratory tract microbiome and lung cancer; (*iii*) the intratumor microbiomes. The third one is is the focus of this study.

Zaneveld *et al* (2017) argued that the microbiome changes induced by many perturbations such as HIV/AIDS infections are stochastic, consequently leading to transitions from stable to unstable community states, or the so termed dysbiosis. The net effects to microbiomes are usually exhibited in the form of an “Anna Karenina principle (AKP)”, in which dysbiotic individuals are more dissimilar in their microbial community compositions than healthy individuals. The AKP was so termed thanks to its parallel to Leo Tolstoy’s aphorism that ‘*all happy families look alike; each unhappy family is unhappy in its own way.*’ Transferred to microbiome-associated diseases, the AKP can be translated as ‘*all healthy microbiomes are similar; each dysbiotic microbiome is dysbiotic in its own way*’ (Zaneveld *et al* 2017). Zaneveld *et al* (2017) inferred that AKP effects should be a common and important response of animal microbiomes to stressors, which lower the ability of the host or its microbiome to regulate (especially stabilize) community composition. The AKP patterns have been observed in systems from the surface of threatened corals, through the lungs of HIV/AIDS patients, to 20+ kinds of human microbiome associated diseases (Zaneveld et al. 2017, Ma 2020b).

The interactions between microbiomes and their hosts are bidirectional. On the one hand, the microbiome and metagenome they carry influence the health, development., disease susceptibility, and even behavior on ecological time scale and influence the host fitness. On the other hand, the host regulates its microbiome. The regulation mechanisms they employ, such as competitions for resources, antibiotic production, metabolic inhibition, spatial occlusion, as well as feedbacks with host immunity, are rather diverse with certain common optimization goals such as preventing invasions by pathogens and managing symbiotic microorganisms to benefit most from them. Zaneveld *et al* (2017) argued that, normally, the defensive activities of hosts and their microbiomes should restrict microbiome membership, avoiding the too diverse detrimental possibilities in favor of a smaller range of beneficial microbiome compositions (configurations). An inference from this argument or observation can be that: stressors, such as cancer disease or HIV infection, need not, at least principally, shift a microbiome to a specific dysbiotic configuration in order to reduce host fitness. Instead, the stressors may only need to release normal regulation of microbiome membership—raising the stochasticity or heterogeneity of community compositions. The stochastic changes often induce dispersion (deviation from some centrality metric such as mean or median), which can be measured with beta-diversity effectively, rather than location (shift of the centrality *per se*).

One of the commonest mechanisms that can induce AKP effects is the compromised host immunity. HIV/AIDS is an example that demonstrated the compromised immunity and AKP effects. There is hypothesis that animal regulation of immunity is evolved to both filter out pathogens and carefully regulate commensal and mutualistic microorganisms. The hypothesis may explain the microbiome instability or dysbiosis in a variety types of immune dysregulation. We further argue that cancers can be considered as typical stressors to host physiology, including their microbiomes and immune systems. Similar to HIV/AIDS infections, cancer cells suppress both host immune system and also host gut microbiome. It has been found that the microbiomes of HIV/AIDS patients follow the AKP (Zaneveld *et al*. 2017, Ma 2020). In particular, Zaneveld *et al*. (2017) presented mechanistic interpretation on why HIV/AIDS impacted microbiomes should follow the AKP. The analysis by Ma (2020b) suggested that the pairwise comparison between HIV-negative (healthy control) and HIV-positive patients followed the AKP, while the comparison between treated and untreated HIV patients followed the anti-AKP. The anti-AKP effects might be due to the treatment (drug) effects that promoted the host immunity.

In summary, the IOM (immunity-oncology-microbiome) axis or perhaps more accurate the IOM trio can exert either positive or negative interaction (feedbacks) to each other within the trio, we postulate that both AKP and anti-AKP effects are possible due to the potential complex feedbacks, similar to the previous HIV/AIDS infections (Fig 1). We further hypothesize that it should be the net effects of the feedbacks within the IOM trio on the host’s regulation capacity to the microbiome composition that matter mostly. When the net effects lead to loosened regulation of the microbiome, then the AKP effects should emerge. Conversely, when the net effects lead to tightened regulation, then the anti-AKP effects should emerge. When the net effects do not significantly impact the regulation, for example, stress or negative impacts from cancer cells may be canceled out by a boost from immune system directly or indirectly (*e.g.,* immune suppression to cancer cells) or from anti-cancer treatment, then no significant AKP effects may be detected.

**Fig 1.**
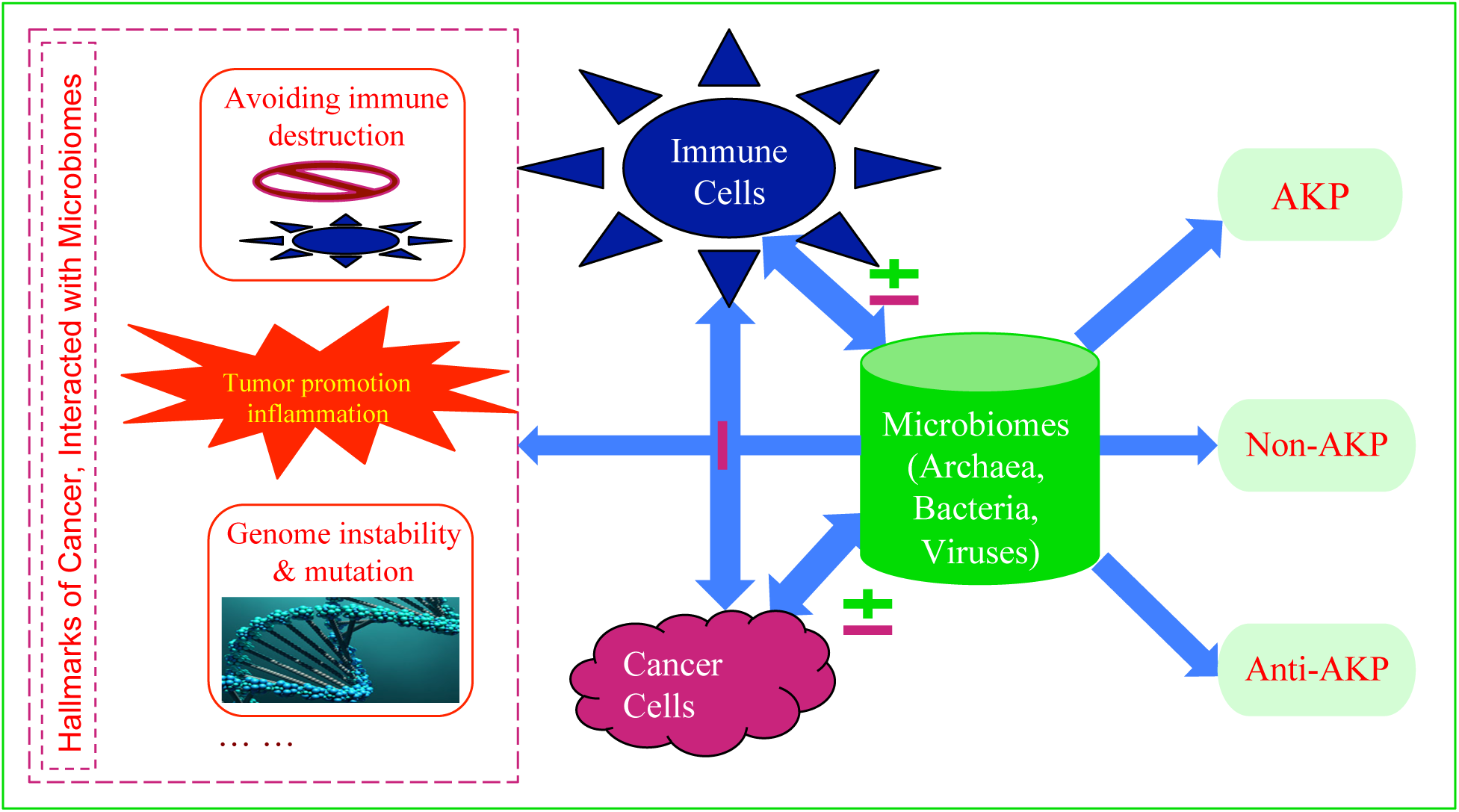
The IOM (Immune-Oncology-Microbiome Axis) axis (paradigm) (Sepich-Poore *et al*. 2021) or the triangle relationships among immune cells, cancer cells and microbes: The left dash-line-framed block shows the three hallmarks of cancer that intersect with the polymorphic microbiomes, which are recognized as a distinctive enabling characteristic that facilitates or hinders the acquisition of hallmark capacities (Hanahan & Weinberg 2011, Hanahan 2022). The right block illustrates our hypothesis that the stresses from cancer cells and immune regulation may generate AKP, anti-AKP or non-AKP effects in the intratumoral microbiomes, depending on the net effects of the “stresses” from cancer and immune cells. Note that all of the interactions between microbes and immune cells or cancer cells are bidirectional. Furthermore, either positive or negative effect on each other, perhaps except for immune-cancer cell interaction (should be antagonistic), is possible, and may even be dynamic (*e.g*., depending on stage of carcinogenesis and/or cancer progression). We further explore whether or not the exhibition of AKP pattern may signal the sign of possible interactions between microbes and cancer cells, *i.e*., whether microbes are oncomicrobes, complicit, bystanders, or foes of cancer cells (see more in Discussion section).

In the remainder of this article, we test the AKP with the intratumoral microbiome datasets of 20+ cancer types, previously extracted from TCGA (the cancer genome atlas) database by Poore *et al* (2020). We further discuss the mechanistic implications of the AKP effects to the IOM, especially the potential roles (oncomicrobes, complicit, bystanders, or foes of carcinogenesis) of intra-tumoral microbes in carcinogenesis and progression, as well as the regulations/perturbation (from the host) that microbiomes have to endure (Fig 1).

## Material and Methods

### Datasets of Cancer Microbiomes

Poore *et al*. (2020) developed a highly sophisticated and powerful bioinformation pipeline, and used it to obtain the microbial (bacteria, archaea and viruses) reads of 33 cancer types from the whole-genome sequencing (WGS) and whole transcriptome sequencing (RNA-Seq) data deposited in TCGA (The Cancer Genome Atlas) database. Their pipeline processed a total of 18116 samples collected from 10481 patients. In this study, we exclude the datasets of 10 cancer types that do not have normal tissue (NT) controls, and reanalyzed the datasets of remaining 21 cancer types, which have primary tumor (PT), solid normal tissue (NT) and blood (B) samples for testing the AKP (Anna Karenina principle). Specifically, we used the RNA-Seq datasets to test the AKP by comparing the PT and NT samples, and WGS datasets by comparing the PT and Blood samples. The NT samples and blood samples are considered as the healthy control samples in both cases. The datasets we reanalyzed (http://cancermicrobiome.ucsd.edu/CancerMicrobiome_DataBrowser/) are available in public domain, and so does the pipeline (https://knightlab.ucsd.edu/) for generating the datasets (see Table S1 for a brief description).

The 21 cancer diseases (types) span six human organ systems, including digestive, urinary, reproductive, central nerve systems. A total of 2117 PT and 1827 blood samples sequenced with WGS protocol, and 11741 PT samples and 641 NT samples with RNA-Seq protocol, respectively are used to perform the AGP tests. The medium of per sample reads was 103, 194. From those PT, NT and Blood samples, the respective OTU tables for archaea, bacteria and viruses were computed in Poore *et al*. (2020), which we reanalyzed for our own analyses for AKP tests. Table S1 exhibited brief summary on the 21 cancer types and their datasets reanalyzed in this study.

### Computational methods for testing the AKP

Zaneveld *et al*. (2017) suggested to use beta-diversity as a metric for detecting potential AKP effects given that rising heterogeneity in community composition is considered as a hallmark of the AKP. There are at least two types of beta-diversity metrics: additive and multiplicative, depending on how the gamma diversity (total diversity of the metacommunity, which consists of 2 or more local communities) is partitioned to measure the difference between local communities, either additively or multiplicatively.

Ma (2020b) applied Hill numbers, which are considered as advantageous over other existing alpha-diversity measures (Ellison 2010, Chao et al. 2012, 2014) and their multiplicative partition is more appropriate than additive partition for defining beta-diversity, for detecting the AKP effects in 20+ microbiome-associated diseases. The Hill numbers are essentially Renyi’s (1961) entropy, of which familiar Shannon’s entropy is a special case. When applied to measure biodiversity, Hill numbers (*H*) are stratified in terms of the so-termed diversity order (*q*=0, 1, 2, …). When *q*=0, *H*=species richness; when *q*=1, H is a function of Shannon’s extropy; when *q*=2, measuring the effective number of species number with typical abundances, when *q*=3, Hill numbers measures the effective number of more abundant or dominant species. Therefore, Hill numbers can offer comprehensive characterizations of community compositions stratified by diversity order (*q*) which represents the shape (skewness) of species abundance distribution (SAD). Obviously, it should be more appropriate to measure community compositions when the shape of is in full consideration.

The beta-diversity in Hill numbers is defined multiplicatively, *i.e*., it is defined as the ratio of the gamma-diversity (total diversity) of metacommunity (consisting of two or more local communities) to the alpha-diversity of local community. For the metacommunity consisting of two local communities (*e.g*., community of PT samples vs. community of NT samples), their beta-diversity ranges from 1 to 2, with 1 meaning that both local communities are the exactly same, and 2 meaning that both local communities are totally different in their compositions. Beta-diversity is also defined with corresponding to diversity order (*q*) to alpha-diversity.

We also applied four similarity measures, which are defined based on beta-diversity and are more convenient for measuring the differences (dissimilarity) between communities, given that they are normalized to the range between 0 and 1. The four similarity metrics are functions or generalizations of the four classic similarity indexes in community ecology, including Jaccard, Sorensen, Horn, and Morisita-Horn measures (Chao *et al*. 2014). They are well tested and recognized as reliable metrics for comparing community compositions, and we use them to cross-verify the results of AKP tests with beta-diversity.

While beta-diversity and four similarity measures are used to quantify the difference between microbiotas of PT *vs*. NT or PT *vs*. Blood samples, non-parametric significance test (Wilcoxon test) is used to determine the statistical significance of the differences in beta-diversity or similarity measure between the healthy (NT or Blood) and diseased (PT) treatments. When the pair-wise beta-diversity within the healthy treatment is significantly lower (higher) than that within diseased treatment (*P*<0.05 from Wilcoxon test), then it is judged that the AKP effects (anti-AKP effects) are detected in the cancer-stressed system. When there is no significant difference in beta-diversity, between the healthy (NT, blood) and diseased samples (PT), no AKP effects are detected. Similarly, the AKP tests can be performed with the four similarity measures. The computational procedures/codes for performing the previously outlined tests are available in Ma (2020b).

## Results

Among the datasets of 33 cancer types published by Poore *et al* (2020) who distilled the microbial OTU tables from the TCGA database, those for the 21 cancer types are suitable for the AKP tests. Among the 21 cancer types, the microbial OTU tables of 11 pairs of NT (solid normal tissue) and PT (primary tumor) treatments are generated from RNA-Seq protocol, those of 19 pairs of Blood and PT treatments are generated from WGS (whole-genome sequencing) protocol. We performed the AKP tests for the RNA-Seq and WGS, separately; in fact, the test is performed for each pair of the healthy (NT or Blood) and diseased (PT) independently. This is, we test the AKP for each of the 21 cancer types, independently, whether it is RNA-Seq or WGS protocol data. Since some of the cancers have both RNA-Seq and WGS datasets, there were a total of 30 AKP tests performed.

Table S2 tabulated the averages of the beta-diversity and the four similarity measures for each treatment (either PT, NT, or Blood) of the 60 treatments (11 pairs or 22 RNA-Seq, + 19 pairs or 38 WGS). Note that those averages for each treatment were obtained from first computing the beta-diversity for each pairs of samples within the focal treatment, and then computing their average. For each pair of treatments, Wilcoxon test was applied to test their differences in beta-diversity and four similarity metrics, respectively.

Table S3 tabulated the *P*-values from the AKP tests for each of the 21 cancer diseases (or 30 AKP tests in total due to the overlap in sequencing protocol), in terms of beta-diversity and each of the four similarity metrics, respectively. That is, 11 sets of tests (with RNA-Seq) and 19 sets of tests (WGS), were performed for the 21 cancer types. Each set of the 30 tests included five Wilcoxon tests based on beta-diversity and the four similarity metrics, respectively. By looking up Table S3, one can find out whether or not each of the 21 cancer diseases is consistent with the AKP, anti-AKP, or none of them. Fig 2 (A-D) (for RNA-Seq) & Fig 3 (A-D) (for WGS) illustrated the test results displayed in Table S3, which we further explain below.

**Fig 2.**
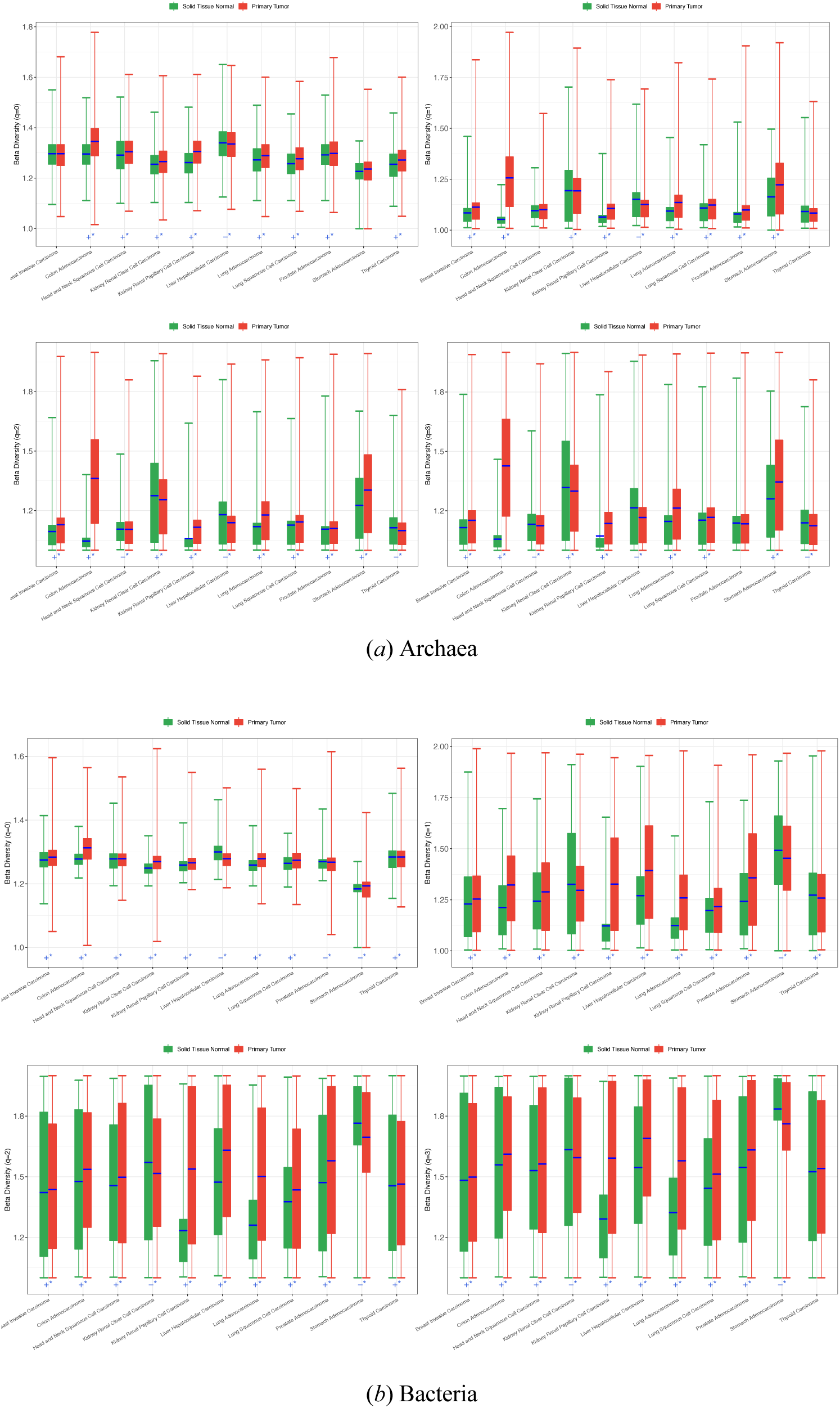
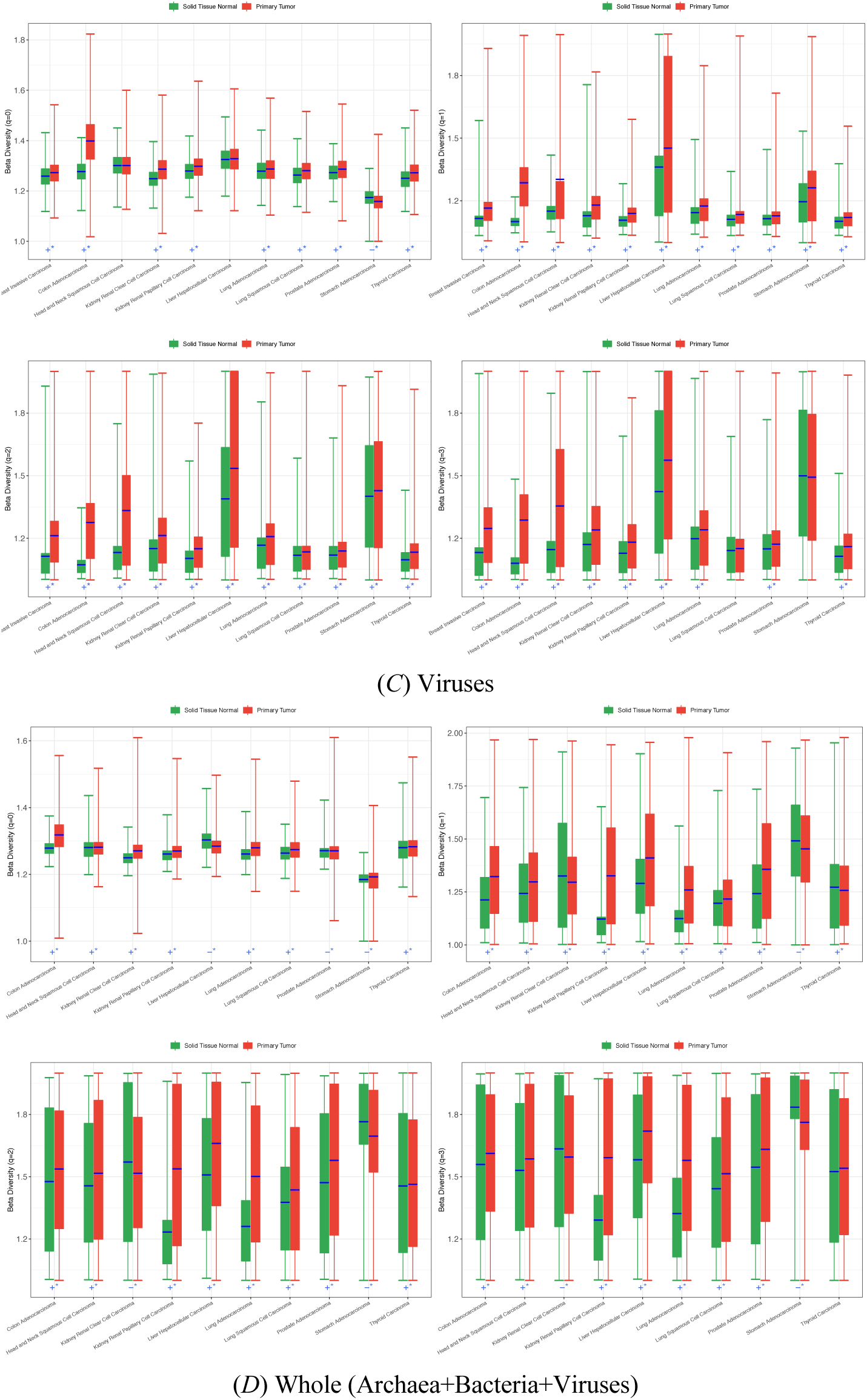
The AKP (‘*+’ for AKP, ‘*–’ for Anti-AKP) tests of 11 cancer types based on the RNA-Seq datasets by comparing the beta-diversity of the NT (solid normal tissue) and PT (primary tumor) microbiomes

**Fig 3.**
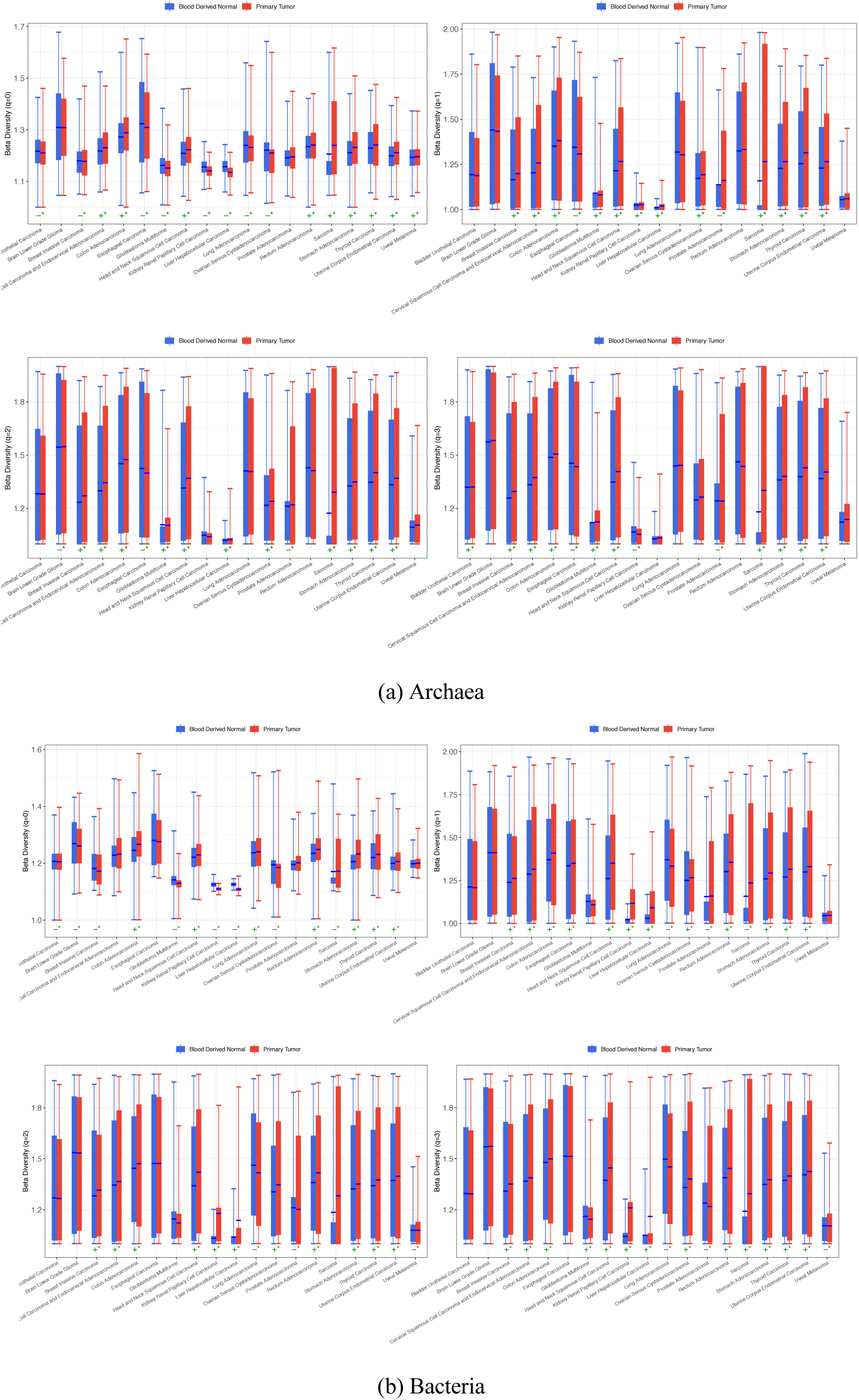
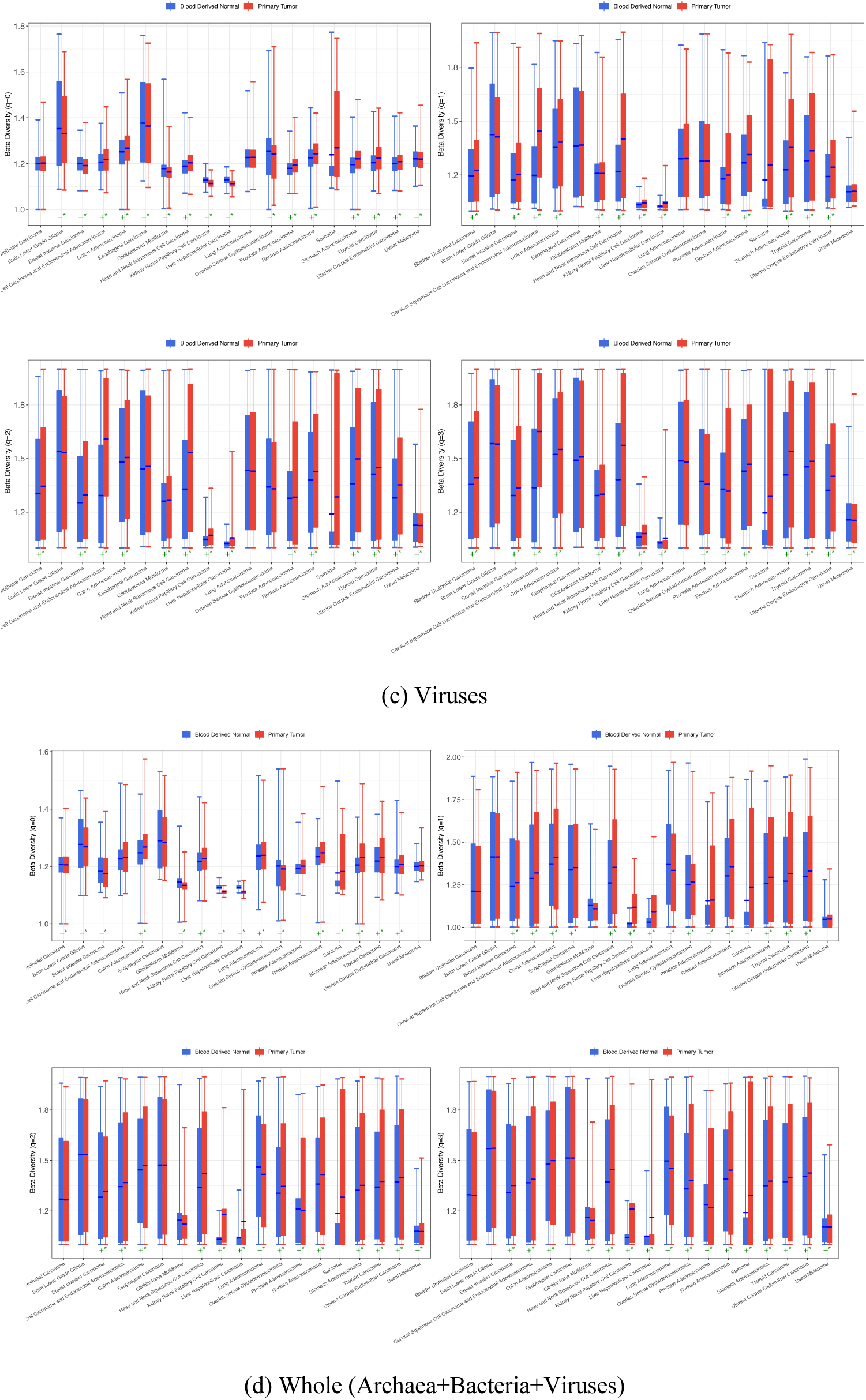
The AKP (‘*+’ for AKP, ‘*–’ for Anti-AKP) tests of 19 cancer types based on the WGS datasets by comparing the beta-diversity of the Blood and PT (primary tumor) microbiomes

Table S4 listed the percentages of cancer diseases that passed (or failed) the AKP tests, based on the beta-diversity, including the percentage of cancer diseases with AKP, anti-AKP, and non-AKP effects, respectively. Table S5 listed the beta-diversity counterpart (percentage) results based on each of the four similarity metrics. Complementary to Table S4, Table S6 exhibited the names of cancer diseases in each of the three categories (AKP, anti-AKP, and non-AKP). Note that the AKP test is performed for archaea, bacteria, viruses, and the whole (all three taxa combined) at each diversity order (*q*=0, 1, 2, 3), respectively.

Fig 2 (A-D) show the graphs of AKP tests (based on Wilcoxon test with beta-diversity) with RNA-Seq data of NT and PT treatment pairs, for each diversity order (*q*=0, 1, 2, 3) and each microbial taxon (archaea, bacteria, viruses, and whole—the combined). In each graph (standard box chart), the *X*-axis is the cancer disease, and *Y*-axis is the beta-diversity for each corresponding cancer disease. Fig 3 (A-D) show the counterpart AKP test results with WGS data of Blood and PT treatment pairs. The box charts show the first quartile (lower edge of the box), the second quartile or the median (the inside segment) and the third quartile (the upper edge) of beta-diversity. The cancers passing AKP and anti-AKP tests are marked with symbols “+*” and “−*”, respectively (no species mark for non-AKP).

Across the 21 cancer diseases (types), Table S4 shows the ranges of percentages of cancer diseases belonging to AKP, anti-AKP, and non-AKP category, for archaea, bacteria, viruses, and their combined (“whole”), respectively. Table S5 shows the counterpart results of the beta-diversity based on four similarity metrics. Figs 3 further illustrated the proportions of AKP, anti-AKP and non-AKP using the “whole” (combined of archaea, bacteria, and viruses) as example. Three patterns can be drawn from Tables S4 & S5 and Fig 4. First, the AKP effects are prevalent, regardless of microbial taxa or sequencing protocols. That is, there are more cancer diseases classified into the AKP category that into anti-AKP or non-AKP category. Second, the proportions of AKP category are higher in the RNA-Seq datasets than in the WGS datasets, which suggests that the expressions of AKP effects are profound in RNA than in WGS datasets and should be expected since WGS datasets may contain genes that are not expressed in transcription processes. Third, the three microbial taxa may have different level of AKP effects in terms of RNA-Seq datasets, with viruses appear having slightly higher AKP percentage than bacteria, and archaea having lowest percentage. However, the differences in taxa seem insignificant in terms of WGS datasets.

**Fig 4.**
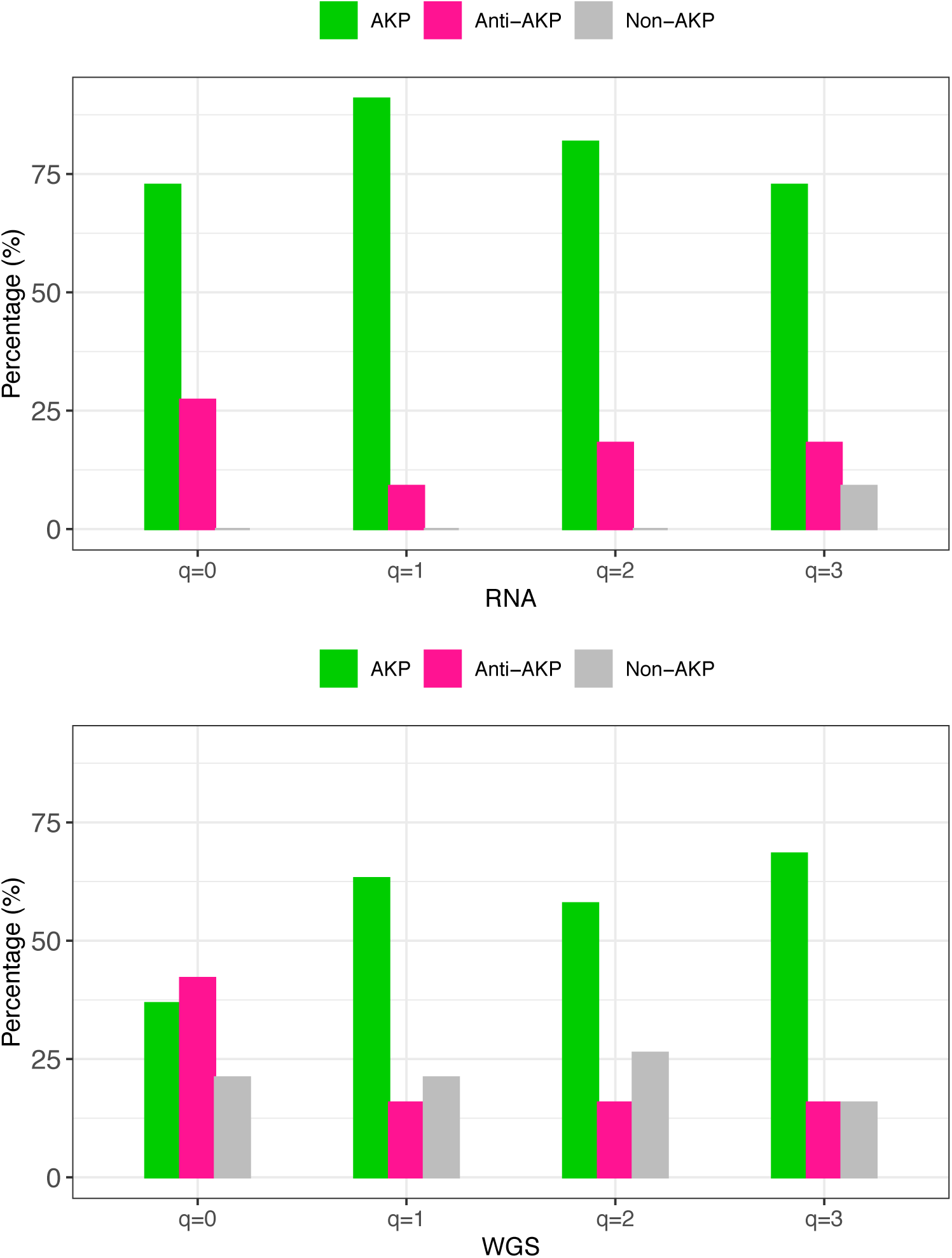
Percentages of AKP, anti-AKP and non-AKP patterns of the “whole” taxa (archaea + bacteria + viruses) at different diversity order (*q*=0-3): the top graph is from RNA-Seq datasets and the bottom is from WGS datasets. When averaged across all diversity orders (*q*=0-3), the ratios of AKP *vs*. anti-AKP *vs*. non-AKP are equal to 9:3:1 for the RNA-Seq datasets, and 2:1:1 for the WGS datasets.

As illustrated in Fig 4, in terms of the “whole” taxa based on RNA-Seq protocol data, the percentage of AKP cancers ranged from 73% to 91%, or approximately ¾ or above; the percentage of anti-AKP cancers ranged from 9% to 27%; the percentage of non-AKP cancers ranged from 0 to 9%. Roughly, the ratio of AKP, anti-AKP, and non-AKP is about 9:3:1; and the ratio of AKP to anti-AKP is about 3:1.

In terms of the “whole” taxa based on WGS protocol data, the percentage of AKP cancers ranged from 37% to 68%; the percentage of anti-AKP cancers ranged from 16% to 42%; the percentage of non-AKP cancers ranged from 16 to 26%. Roughly, the ratio of AKP, anti-AKP, and non-AKP is about 2:1:1; and the ratio of AKP to anti-AKP is about 2:1. Obviously, as mentioned previously, the AKP effects are more profound than Anti-AKP effects in the RNA-Seq datasets, that is, the genes responsible for AKP effects are expressed more profoundly than the genes responsible for anti-AKP effects during transcription processes. This seems to vindicate that the AKP effects are indeed induced by cancer stresses.

## Conclusions and Discussion

The previous AKP tests of 21 cancer types (Poore *et al*. 2020), including 11 tests with RNA-Seq (transcriptome) datasets and 19 tests with WGS (whole genome) datasets, based on beta-diversity and four similarity metrics (Ma 2020b) reveal the following findings: First, the AKP is prevalent over anti-AKP and non-AKP, especially with transcriptome datasets, the ratios of AKP *vs*. anti-AKP *vs*. non-AKP are 9: 3: 1 in RNA-Seq datasets, and the ratios are 2: 1: 1 in the WGS datasets. Ignoring non-AKP, the ratio of AKP to anti-AKP is 3:1 and 2:1 for the RNA-Seq and WGS, respectively. The higher prevalence of AKP in RNA-Seq than in WGS datasets should be expected since it simply reflects the fact that RNA-Seq protocol only sequenced expressed or transcriptomed functional genes, while the WGS protocol sequenced both expressed and non-expressed genes. The non-expressed genes should be stable (homogenous) across the healthy control (blood or NT) and PT samples, and consequently of lower AKP effects. Second, With WGS data, the differences among taxa in their AKP patterns seem negligible. With RNA-Seq data, viruses appear to exhibit the highest AKP effects, followed by bacteria and the archaea exhibiting the lowest. Accordingly, viruses exhibited the lowest anti-AKP effects. We could not perform statistical tests for the taxon-level differences with currently available datasets, and the differences could be insignificant.

As introduced previously, current investigations on the relationships between human microbiomes, immune cells and cancer cells or the IOM (immune-oncology-microbiome) have been focused on three areas: the gut or intestinal microbiome, non-intestinal barrier organ microbiomes (oral, skin, respiratory tract, vaginal) and intratumoral or tumor-tissue microbiomes. In the remainder of this article, we briefly summarize the three areas, heavily drawn from excellent reviews by Sepich-Poore *et al* (2021) and Hanahan (2022), in order to infer possible mechanistic insights from our AKP tests in the context of IOM, or the trio of cancers, microbiomes and immune systems. Below, we first summarize the three areas in three respective paragraphs, and then discuss our inferences and hypothesis.

Since the gut offers the largest host-microbial interface and highest microbial diversity, there is no surprise that investigations on the potential impact of the microbiota on oncogenesis and cancer prognosis have revealed most findings to date on the relationships between gut microbiome alternations and compromised tumor immuno-surveillance, including extraintestinal cancers. The high density of colonic bacteria is considered to be the most significant driving force of microbial immunomodulatory effects in the mammalian intestinal tract, which is considered the largest immune organ in our body. A key aspect of the microbiome-cancer relationship is then naturally the mechanisms and interactions between the gut microbiome and cancer development, given that gut microbiome can regulate many functions of the tumor-bearing meta-organism or holobiont, typically via immunomodulation. Existing mechanistic and interactions studies on gut microbiomes have been focused on the effects of gut microbiota on primary lymphoid organs, adaptive immunity, and TME, as well as on their mediation on anticancer drugs. Based on such background studies, Sepich-Poore *et al* (2021) defined the immune-medicated interactions and collective feedback loops as the immune-oncology-microbiome (IOM) axis.

Beyond the gut microbiome, the importance of local microbiomes in extraretinal cancers cannot be denied, and may play a role in the exacerbation of neoplasia (Jin *et al*. 2019; Le Noci *et al*. 2018; Nejman *et al* 2020; Tsay *et al* 2020). For example, the lung’s surface, with approximately *1* m^2^ per kg of body weight, hosts its own local microbiota. The local commensals could be disturbed by carcinogenesis, inducing an inflammatory cross-talk between alveolar macrophages and IL-17-producing lung resident *γδ* T cells contributing to tumor progression (Jin *et al*. 2019; Tsay *et al* 2020). Skin microbiome is also important for maintain host homeostasis through its interactions with skin immune components including metabolic, innate, and cognate immune responses (Stacy *et al*. 2019). Compositional changes in the skin microbiome seems implicated in the nonmelanoma skin carcinogenesis (Squarzanti *et al* 2020).

In spite of the obvious putative importance of intratumoral microbiome and organ-specific commensals, current understanding of intratumoral microbiomes is poorly understood at present. The abundance of intratumoral microbiome is quite low. Nejman *et al* (2020) estimates suggested an average pan-cancer percentage bacteria composition at 0.68% bacterial, which could be converted to ∼10^5^ to 10^6^ bacteria per palpable 1-cm^3^ tumor or ∼34 bacteria/mm^2^ (assuming 5000 cells/mm^2^) (Sepich-Poore *et al*. 2021). This latter number is close to the mean tumor core density of ∼21 cells/mm^2^ from a recent pan-cancer cohort (Cited in Sepich-Poore *et al*. 2021). Mechanistic studies on intratumoral microbiomes have been rather limited, except for the aerodigestive tract. Several studies have identified their suppressive effects on local antitumor immunity via the TME (Hamada *et al*. 2018; Le Noci *et al*. 2018; Mima *et al*. 2015; Pushalkar *et al*. 2018; Parhi *et al*. 2020; Riquelme *et al*. 2019; Roberti *et al*. 2020). In addition, there have been some reports on the cancer-specific effects of intratumoral microbiomes. Sepich-Poore *et al* (2021) reviewed five categories of those cancer-specific influences, including: (*i*) gastrointestinal and urinary tract mutagenesis through secreted genotoxins (Barrett *et al*. 2020; Boot *et al*. 2020; Dejea *et al*. 2018; Pleguezuelos-Manzano *et al*. 2020; Wilson *et al*. 2019), (*ii*) CagA-mediated inflammation in stomach cancer (Jin *et al*. 2019) or IL-17–producing *γδ* T cell–mediated inflammation in lung cancer (Suzuki *et al*. 2009), (*iii*) chemoresistance via direct microbial metabolism in pancreatic cancer (Geller *et al*. 2017) or indirect amplification of cancer cell autophagy in colorectal cancer (Yu *et al*. 2017), (*iv*) tumor proliferation via fungal activation of pancreatic cancer patient’s C3 complement cascade (Aykut *et al*. 2019); (*v*) metastasis via increasing tumor matrix metalloproteinases in breast cancer (Parhi *et al*. 2020) or lowering tumor immunosurveillance in lung cancer (Le Noci *et al*. 2018). Intratumoral microbiomes often generate tolerogenic programming and produce immunological tolerances, but there are exceptions as demonstrated by the injection of intratumoral bacteria or their antigens could conversely stimulate immune responses. These exemplary studies appear to suggest that intratumoral microbes are more likely to be complicit of cancer cells, rather than their adversaries. Nevertheless, the associations between intratumoral microbiomes and immunogenicity were also discovered. That is, intratumor microbes may stimulate immune response to cancer cells. In summary, the net effects of the intratumoral microbiomes, similar to the gut and other barrier-organ (skin, lung, *etc*.) microbiomes, can be either protective or deleterious effects on cancer development, malignant progression, and therapy responses. Of course, microbiomes have to endure the “anti-force” or perturbations from cancer cells and modulation from immune cells. The effects of the perturbations/modulation are exhibited in the form of AKP, anti-AKP, or non-AKP on ecological time scale; nevertheless, an evolutionary perspective is necessary to shed mechanistic insights on the patterns of AKP, as elaborated below.

After briefly summarizing the state-of-the-art studies on the IOM or the trio of immune cells, cancer cells and microbes in previous paragraphs, we further discuss the potential implications of the AKP effects in the context of the IOM or the trio, from a unified evolutionary and ecological perspective. Inspired by Hutchinson’s (1957) classic “The ecological theater and evolutionary play,” cancer can be considered as an evolving ecosystem consisting of tumor cells, stem cells, stromal cells, immune cells, microbes, and tumor microenvironment (TME). Cancer evolutions are on multiple levels ranging from ultra-micro (cell population level), micro (individual host) and macro (population of individuals), and can be real time evolution at the ultra-microscale (Horning 2017, Reynolds *et al*. 2020, Dujon *et al*. 2021, Ma & Zhang 2022). What Hutchinson’s (1957) title transpired is that evolution (“play”) depend on ecological environment (“theater”), and the evolution of cancer cell population depends on TME, which includes microbiomes and immune cells. Therefore, we argue that the AKP effects can be considered as exhibitions of eco-evolutionary dynamics of cancer ecosystem, and we further postulate that the AKP effects can also be considered as exhibitions of the IOM axis or the trio of immune, cancer cells and microbes.

The evolutions of metazoans (animals and plants) are inseparable with microbial evolution. Animals are evolved or even originated in a sea of microbes, which prompts some scholars to apply coevolution theory to interpret the symbiotic relationships between microbes and their hosts animals. Some scholars even argue for the genetics and evolution of holobiont (host + their symbiotic microbiomes) and hologenome carried by the holobiont (Rosenberg *et al*, 2018, Simon *et al*. 2019, Ma 2021, Ma *et al*. 2022). Since microbes existed and evolved well before the emergency of multicellular animal and plants, the evolution of animals and plants should have evolved to defense against exploitation of opportunistic microbial pathogens and to benefit from microbial metabolic innovations via mutualistic microbiomes (Zaneveld *et al*. 2017). The associations between microbiome dysbiosis (instability) and many stresses/diseases such as cancer and HIV hint that microbiome stability (resistance and resilience) are a hallmark of healthy physiology, which is consistent with the evolutionary history of animals, *i.e*., animals are evolved in a sea of microbes. The AKP tests provide an effective approach to measuring the effects of dysbiosis by comparing to normal variations in healthy microbiomes with statistical rigor, as performed previously with the datasets from TCGA database.

While a primary take-home message of this study is that stresses from cancer cells and modulation from immune cells can lead to rising (AKP) or declining (anti-AKP) inter-subject (inter-host) heterogeneity of intratumoral microbiomes, depending on the net effects of the stresses/modulation, we further ask whether or not observed AKP effects may reveal the underlying mechanistic interactions (within the IOM) that generated the observed patterns. The emergence of AKP may suggest that the relationship between majority members of the microbiome and cancer cells should be more stressful (contentious) than their random interactions. Conversely, the emergence of anti-AKP may suggest that the relationship should be less stressful or even complicit than their random interactions. The lack of AKP effects (non-AKP) may suggest that the interactions between cancer cells and intratumoral microbes should just be random. Nevertheless, the “stressful” (contentious) relationship between intratumoral microbiomes and cancer cells do not necessarily mean that their direct interactions are negative due to the existence of the third party—immune cells. That is, the immune system may alter the balance between cancer cells and intratumoral microbes. In other words, whether or not some intratumoral microbes are oncomicrobes (human carcinogens), complicit, bystanders, or foes do not necessarily determine the AKP effects are positive (AKP), negative (anti-AKP) or null (non-AKP) because of the modulation of immune system. Indeed, as Hanahan (2022) suggested that the effects of microbiome on cancer cells could be intersected with immune system.

What if the immune system effects on microbes and cancer cells are constant? This is not necessarily an unrealistic assumption at some stages of cancer development. Even if this assumption is satisfied, there is still uncertainty, which may prevent us from inferring the mechanistic interactions between cancer cells and intratumoral microbes. The uncertainty is that the AKP effects are largely a microbiota-level or holistic characteristic, and individual or even some cancer complicit or foes do not necessarily have sufficient power to sabotage the holistic characteristics. Nevertheless, when their effects are sufficiently strong to lead to dysbiosis (instability) of microbiota, the AKP effects should be able to reveal the mechanistic nature of the interactions between cancer cells and intratumoral microbes, *e.g*., oncomicrobe, complicit, foe, or bystanders.

Despite the limited value of AKP effects in revealing the mechanistic interactions between cancer cells and intratumoral microbes, the detection of AKP effects is still of important biomedical significance. From biomedical research perspective, as mentioned previously, predicting the response of microbiome to perturbations including diseases is challenging but rewarding. Investigating the responses in the presence of cancer treatment may also offer additional insights on personalized treatment of cancer. From a pure ecological research perspective, to the best of our knowledge, the AKP tests we adopted in this study (Ma 2020b) are far more sensitive than other community-level metrics such as alpha-diversity, which usually only detect approximately 1/3 of the changes associated with diseases (*e.g*., Ma *et al*. 2019, Ma & Li 2019, Ma & Ellison 2019, Ma 2020a, 2020c), while the total of AKP and anti-AKP are close to 90%. A limitation of AKP tests is its lack of consideration of individual microbial species, but this can be addressed by complementary approaches such as Ma & Li (2022) & Ma (2022), which can detect unique or enriched species in cancer cells. A preliminary result from the integration of both AKP tests and the unique/enriched species detection (Ma & Li 2022) is that, in general, the unique species are significantly more in PT (primary tumor) than in NT or blood microbiomes. In some cancers, there is not any unique species in healthy microbiomes, which highlights the undeniable effects of cancer in altering microbiome composition (Ma & Li 2022).

## Data Availability

Availability of datasets
The datasets reanalyzed in this article is publicly available from the Online Supplementary Information of the following publication: Poore, GD, E Kopylova, Q Zhu, R Knight (2020). Microbiome analyses of blood and tissues suggest cancer diagnostic approach. Nature 579, 567-574. https://doi.org/10.1038/s41586-020-2095-1

## Online Supplementary Information

**Table S1**. Brief summary on the microbial datasets distilled from the TCGA (the cancer genome atlas) by Poore et al (2020) and reanalyzed for the AKP tests

**Table S2**. The mean of beta-diversity and four similarity indices for the 21 cancer diseases (MS-Excel File)

**Table S3**. The P-value of the Wilcoxon test between healthy site and diseased site for 21 cancer types (MS-Excel File)

**Table S4.** The percentages of AKP, anti-AKP and non-AKP determined from comparing the healthy control (Blood or NT treatments) and primary tumor (PT) in 21 cancer types, based on their beta-diversities (summarized from Table S3)

**Table S5.** The percentages of AKP, anti-AKP and non-AKP determined from comparing the healthy control (Blood or NT treatments) and primary tumor (PT) in 21 cancer types, based on the *four similarity indices* (summarized from Table S3)

**Table S6**. The list of cancers following the AKP, Anti-AKP and Non-AKP patterns, respectively, summarized from Table S3 (note that the test results from beta-diversity and four similarity metrics are the exactly same).

## Acknowledgements

I appreciate the computational support from Dr. Lianwei Li of Chinese Academy of Science. This study received funding from the following sources: A National Natural Science Foundation (NSFC) Grants (No. 31970116, 72274192).

## Availability of datasets

The datasets reanalyzed in this article is publicly available from the Online Supplementary Information of the following publication: Poore, GD, E Kopylova, Q Zhu, … R Knight (2020). Microbiome analyses of blood and tissues suggest cancer diagnostic approach. *Nature* 579, 567–574. https://doi.org/10.1038/s41586-020-2095-1

## Author contributions

ZSM designed the methodology and analysis scheme, wrote the manuscript.

## Ethic Approval

N/A since the study does not involve any wet-lab experiments or survey on human or animal subjects, and all analyzed datasets are already available in public domain, as mentioned above.

## Conflict of interests

The author declares no conflict interests.

## Notes

### Competing Interest Statement

The authors have declared no competing interest.

